# Simulation-based Estimation of the Spread of COVID-19 in Iran

**DOI:** 10.1101/2020.03.22.20040956

**Authors:** Navid Ghaffarzadegan, Hazhir Rahmandad

**Affiliations:** Virginia Tech, Department of Industrial and Systems Engineering; MIT, Sloan School of Management

## Abstract

**Background:** The COVID-19 disease has turned into a global pandemic with unprecedented challenges for the global community. Understanding the state of the disease and planning for future trajectories relies heavily on data on the spread and mortality. Yet official data coming from various countries are highly unreliable: symptoms similar to common cold in majority of cases and limited screening resources and delayed testing procedures may contribute to under-estimation of the burden of disease. Anecdotal and more limited data are available, but few have systematically combined those with official statistics into a coherent view of the epidemic. This study is a modeling-in-real-time of the emerging outbreak for understanding the state of the disease. Our focus is on the case of the spread of disease in Iran, as one of the epicenters of the disease in the first months of 2020.

**Method:** We develop a simple dynamic model of the epidemic to provide a more reliable picture of the state of the disease based on existing data. Building on the generic SEIR (Susceptible, Exposed, Infected, and Recovered) framework we incorporate two behavioral and logistical considerations. First we capture the endogenous changes in *contact rate* (average contact per person) as more death are reported. As a result the reproduction number changes endogenously in the model. Second we differentiate reported and true cases by including simple formulations for how only a fraction of cases might be diagnosed, and how that fraction changes in response to epidemic’s progression. In estimating the model we use both the official data as well as the discovered infected travelers and unofficial medical community estimates and triangulate these sources to build a more complete picture. Calibration is completed by forming a likelihood function for observing the actual time series data conditional on model parameters, and conducting a Markov Chain Monte Carlo simulations. The model is used to estimate current “true” cases of infection and death. We analyze the future trajectory of the disease under six conditions related to the seasonal effects and policy measures targeting social distancing.

**Findings:** The model closely replicates the past data but also shows the true number of cases is likely far larger. We estimate about 493,000 current infected cases (90% CI: 271K-810K) as of March 20th, 2020. Our estimate for cumulative cases of infection until that date is 916,000 (90% CI: 508K, 1.5M), and for total death is 15,485 (90% CI: 8.4K, 25.8K). These numbers are significantly (more than one order of magnitude) higher than official statistics. The trajectory of the epidemic until the end of June could take various paths depending on the impact of seasonality and policies targeting social distancing. In the most optimistic scenario for seasonal effects, depending on policy measures, 1.6 million Iranians (90% CI: 0.9M-2.6M) are likely to get infected, and death toll will reach about 58,000 cases (90% CI: 32K-97K), while in the more pessimistic scenarios, death toll may exceed 103,000 cases (90% CI: 56K-172K).

**Implication:** Our results suggest that the number of cases and deaths may be over an order of magnitude larger than official statistics in Iran. Absent extended testing capacity other countries may face a significant under-count of existing cases and thus be caught off guard about the actual toll of the epidemic.

## 1. Introduction

The 2019 novel Coronavirus (SARS-Cov-2), the pathogen that causes COVID-19 infection, is exposing the world to one of its largest global health challenges of recent time. The challenge is multi-faceted, from understanding a quickly evolving situation to responding with limited information and under extreme pressures on the healthcare system. From public understanding to policy choices, much depends on the data about the epidemics spread and models that integrate such data into actionable policies (Kaplan, Craft et al. 2002, Thompson, Tebbens et al. 2008). Yet the official data is highly uncertain, with large variations in quality depending on the country reporting it, and regularly offering lower bounds that have unknown error bounds compared to the reality on the ground. Other sources of data, often based on smaller samples, travel screening, and anecdotal evidence may offer relevant hints, but there are not easily generalized or combined with official data. The current paper focuses on using a standard dynamic epidemiological model as a tool for incorporating various sources of data into a unified estimation of the trajectory of disease for the country of Iran.

### 1.1. Background and Motivation

Current information (as of mid-March 2020) point to the spread of COVID-19 starting from a food market in Wuhan China in mid-November 2019. The epidemic was not officially detected until the early 2020, and spread rapidly, mostly in China, for January and February of 2020. In response significant public health resources were mobilized in China, entire cities and regions were put under quarantine, and economic activity was reduced to a fraction. By mid-February the speed of contagion was slowing down in China, offering hope to many that the epidemic would be contained before becoming a global pandemic. Since then, however, rapid rise of detected cases in many other countries have dimmed the hopes for containing the contagion at origins. The situation may also be much worse than official statistics portray. For example, in late February, the United States had only a handful of endogenous cases, i.e. identified as not being imported from China. Yet genetic sequencing of the virus among one of those cases tied it back to a traveler from China who entered the country six weeks before and was thought to have not infected anybody (Fink and Baker 2020). This implies that the epidemic might have been spreading in the U.S. for six weeks without being detected. Cases of death from COVID-19 might have also been popping up, but attributed to flu instead. The situation may be worse elsewhere. Among countries in south and east Asia with significant economic and travel ties to China, several, including India, Pakistan, Indonesia, Malaysia, Vietnam, Myanmar, Bangladesh, and Philippines have reported limited cases (as of early March) despite having a total population of over two billion. That seems unlikely when many smaller countries or those with further ties from China already show sustained epidemics, unless temperature plays an important role in infectivity, a possibility that requires further investigation (Wang, Jiang et al. 2020). Thus official statistics are not a reliable gauge for understanding the actual extent of the epidemic.

The situation in Iran, the country with the third largest number of official deaths from the disease by late March 2020, is instructive. The first official cases were reported on February 19th, 2020, in the city of Qom (Wikipedia 2020). Later some anonymous sources reported on observing cases suspected of the disease much earlier, as early as January 2020 (IranInternational 2020). Yet in the absence of test kits to confirm the virus and lagging government response, those reports gained little traction. From February 19^th^ on the disease spread rapidly in different cities. How far will this epidemic go and what policies should be put in place to control and mitigate the risks very much depends on understanding the true magnitude of the epidemic.

While large and alarming, current reports for Iran, and many other countries, may well be underestimating the actual size of the pandemic. Any under-estimation is worrisome, but early in the exponential growth phase of an epidemic such errors could be extremely costly. The risk of under-estimation is partly driven by the characteristics of the COVID-19 such as a potentially large population of unrecognized patients with mild symptoms (at least 80% of the cases have symptoms not very different from common cold or flu) (Novel Coronavirus Pneumonia Emergency Response Epidemiology 2020). Some heuristics are used to correct for such biases. For example the ratio of deaths to those recovered at the beginning of March was 6.3% (Worldmeters 2020), yet most reports put the death rate at lower rates (Fauci, Lane et al. 2020, Wu and McGoogan 2020), implying that some 70% of cases go undetected. A formal study puts that number at 86% for the early stage of the epidemic in China (Li, Pei et al. 2020). Yet, the country-specific biases in measurement and reporting may exceed those due to the nature of the disease. To illustrate, in the next section we provide a quick survey of various clues related to the magnitude of epidemic in Iran, which we then build on in our analysis.

### 1.2. Clues to the magnitude of Iran’s epidemic

There have been a few clues from Iran which may inform efforts to estimate the true cases. First, is the number of cases identified among travelers arriving in other countries from Iran. Screening of passengers from high-risk countries in airports is more reliable than most country-level screening statistics. One study in pre-print estimated 18,300 total cases of infected individuals in Iran by February 25 (Tuite, Bogoch et al. 2020). The method relied on estimation of cases in the whole country based on three diagnosed cases of infection upon individuals’ arrival from Iran in various international airports and the likelihood of such an incident given approximately 7,500 daily outbound passengers from Iran during those early days (Fraser, Donnelly et al. 2009). There is much uncertainty associated with these estimates, a wide 95% confidence interval of 3,770 –53,470, partly because the method assumes many cases of infected travelers have gone undetected. Nevertheless the range was orders of magnitude larger than official statistics at the time. Later reports on the number of infected travelers from Iran rose rapidly, to 97 cases by February 28 (RadioFarda 2020). An article in *The Atlantic*, offered a series of back-of-the-envelope calculations which included several simplifying assumptions, estimating 2 million accumulated cases of infected individuals by March 9^th^ (Wood 2020).

One may expect the death statistics to be more reliable. But test kits for identifying COVID-19 have been in short supply, and post-mortem testing may not have been a priority of officials in Iran, so potentially many cases are missed. For example on March 1st a health official in Golestan, a state with 1.9 million population in Iran, reported 594 cases of Coronavirus in the state based on CT-scan outcomes (IranInternational 2020). He complained the cases were not yet counted in the official tally because the central authorities had not provided test kits to the state and are reluctant to accept other diagnosis methods. A BBC Persian report on February 28^th^ used interviews with an unspecified number of hospitals in Iran to put the death from the disease at 210, an order of magnitude larger than official numbers at the time (BBCPersian 2020). Another news agency quoted similar sources for a total of 416 deaths by March 1^st^ (IranInternational 2020) and 5000 on March 18^th^. On February 24^th^ a member of Iran’s parliament reported 50 deaths only in Qom, a city of 1.2 million (Wikipedia 2020). Several reports of government officials contracting the disease have also been released, including reports on infection of 20 member of the Iranian parliament (a body of 270 members), as well as several deaths among officials (BBCPersian 2020). More informal observations shared on social media offer a peek into grim conditions in hospitals, with large numbers of patients suffering from COVID-19 symptoms and death rates that are far larger than official statistics suggest and include many deaths at homes.

### 1.3 Reconciling various clues using a dynamic model

We develop a dynamic simulation model of the spread of the disease in Iran to estimate the likely trajectory of the disease that is consistent with the evidence summarized above. We start with the traditional SEIR (for Susceptible, Exposed, Infectious, and Recovered stocks representing population groups) model and incorporate feedbacks regulating endogenous changes in contact rate, screening, diagnosis, and reporting in response to risk perception and other relevant factors. Thus not only reported statistics, but also the effective reproduction number, R_e_, are endogenously generated and can change as people respond to the epidemic. The model is very simple on other fronts: it assumes perfect mixing for the whole population of Iran and includes no disaggregation of population into different groups, nor any travel patterns across population groups. The focus being on a single country, the only link with the pandemic is when the cases were first seeded in Iran (supposedly from China). We use this model, along with various strands of data reported above, to weave together an estimate of disease trajectory so far, and offer projections for expected future trajectory. Given the rapid development of this model and various uncertainties not included in the analysis, the results should be seen as an assessment to enhance the overall picture of the epidemic, but not as reliable point estimates. With these limitations in mind, we estimate over 916,000 (90% CI: 508K, 1.5M) cumulative cases of the disease in Iran as of March 20th, with over 15,485 (90% CI: 8.4K, 25.8K) deaths. We thus estimate that only 2.1% of cases and 9.2% of deaths are officially attributed to COVID-19, with the rest going undetected. The confidence intervals around our estimations are relatively wide due to data limitations and our conservative assumptions in estimating those intervals. Nevertheless these results point to extreme gaps between official data and actual trajectory of disease, which may lead to slow response and under-appreciation of risks of the diseases in the coming months.

## 2. Methods

### Model Structure

Figure 1 offers a simple representation of the model’s structure. Model equations, and parameter values are documented in the supplementary material. The model belongs to the family of the infectious disease models knowns as SEIR (Susceptible, Exposed, Infectious, and Recovered). Main state variables of the model are shown in the Figure as boxes, with flows between them represented explicitly. The infected population is first asymptomatic and later becomes symptomatic. The inflow to the asymptomatic population mostly comes from the susceptible population. First cases of infection are injected to the exposed stock which then trigger dynamics of infection. These first cases are critical for starting the epidemic, but once several seeds of infection are planted, later importation of cases in largely inconsequential to the overall trajectory (Chinazzi, Davis et al. 2020). The symptomatic infected population will follow two different paths of recovery or death. In practice different symptomatic subpopulations have different risks both in severity of illness and mortality, but to avoid proliferation of parameters we use general population averages.

**Figure-1:**
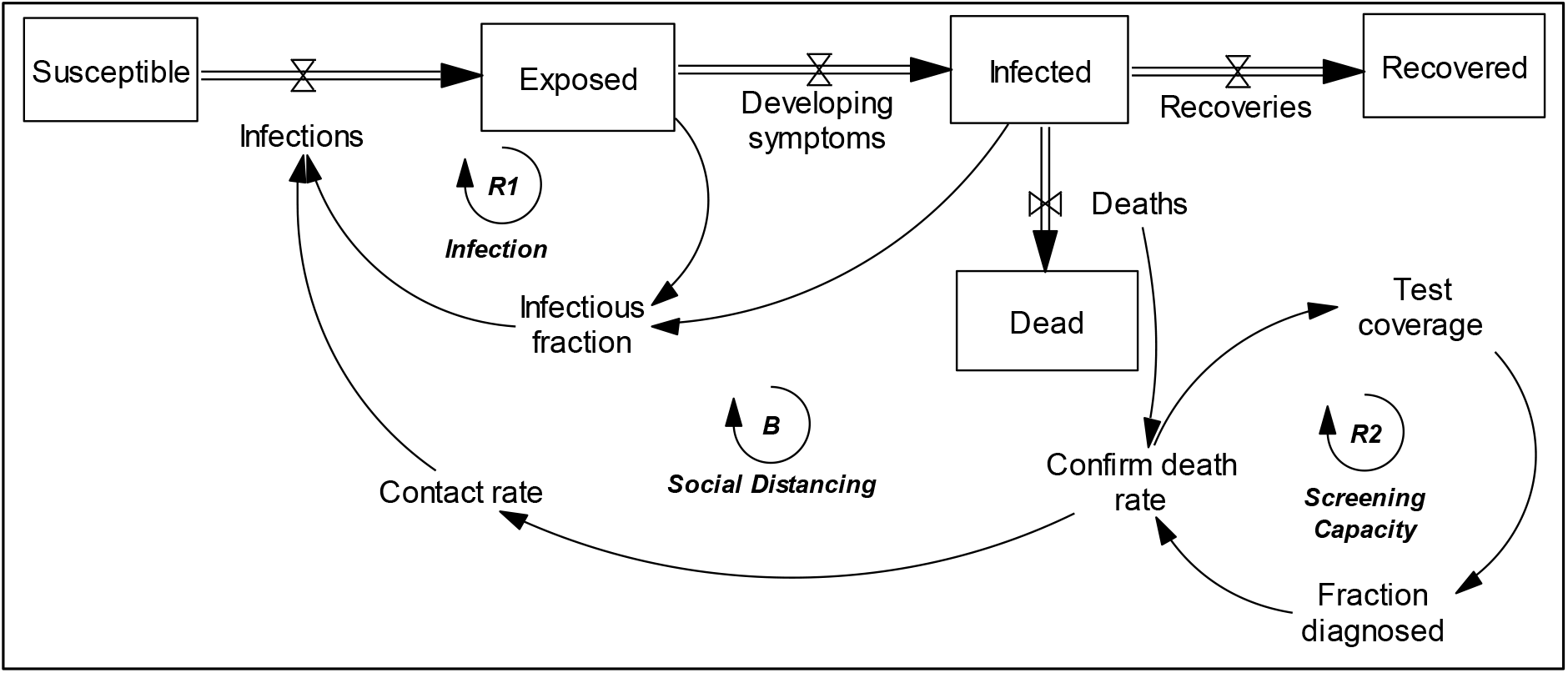
A simplified representation of the model

The model includes the ‘Infection’ reinforcing loop (R1) which regulates the spread of disease from infected to susceptible, and creates the initial exponential growth in the number of cases. We also assume that asymptomatic patients (captured in the ‘Exposed’ stock in Figure 1) might be infective (Bai, Yao et al. 2020), though at a lower infectivity than symptomatic infected. As in the generic SEIR model the disease will continue as long as the reproduction number, the number of secondary cases from each infection, remains above one.

To this basic epidemiological model we add two endogenous mechanisms that are key to understanding the observed trajectories in official data. First, we formulate contact rate to be endogenously changing in response to perceived risk of infection (a function of death statistics). The impact of perceived risk on contact rate captures not only the endogenous changes in social interactions, gatherings, self-isolation of suspected cases, and hygiene, but also government mandated closure of events, schools, and businesses to name a few. For example in Iran schools and sport and cultural events were closed down after the seriousness of the epidemic was recognized in late February. Moreover people started to reduce their discretionary interactions in public spaces, cut down on social and family gatherings, and followed hand washing and mask wearing recommendations more closely. We assume public risk perception depends on the number of recent reported cases of death. This balancing feedback (loop B) can bring down contact rate, potentially enough to slow down the epidemic, and captures in a simple formulation how a few other countries, namely China and South Korea, have brought down their case counts after the initial exponential growth. Second, we explicitly model the endogenous changes in screening and reporting of cases over time. Here, the increased understanding of risks leads to mobilization of screening resources that further expand the case detection and official statistics in a reinforcing process (loop R2). Nevertheless even with good screening many mild cases will go undiagnosed and not counted in official statistics.

### Estimation

We have time series data for official reports of death, recovered, and cumulative number of infection over time. Besides the official data, we use a few unofficial data points including three observations about number of Iranian passengers diagnosed with COVID-19 upon arrival in international airports, and three unofficial estimations from BBC and IranInetrnational news sources about total cases of death from COVID 19. The model includes several biological parameters (such as the asymptomatic period of the disease or average time to recover) which we specify based on prior literature; population size and travel scope are also input using existing data. Eleven uncertain parameters remain that are estimated using the above data. Five of the parameters are used to specify how official measurement and reporting relates to “true” values of infection and death, three parameters estimate public reaction to the reports, and two parameters are for mortality rate among two different groups of patients.

Finally, the arrival of first cases of the virus is estimated as a separate parameter.

Our calibration method is mainly based on forming a likelihood function for observing the actual time series data conditional on model parameters. We then conduct a Markov Chain Monte Carlo (MCMC) simulation to estimate the joint posterior distribution of the model parameters subject to observed data. We define a likelihood function for change over time (net-flow) of official reports on death, recovered and infection assuming they are count events drawn from model-predicted rates (Poisson distribution). We use a similar Poisson distribution assumption for number of infected passengers and unofficial reports of death as well, since they both fit well into a count measure framework. The MCMC method searches over the feasible ranges for the uncertain parameters and accepts various combinations of parameters that are consistent with the observed data. Similar methods are used frequently in estimating dynamic models of epidemics. For example Wu and colleagues used a similar likelihood based MCMC method for estimating the basic parameters of the COVID-19 epidemic and its potential for spread beyond China (Li, Pei et al. 2020, Wu, Leung et al. 2020). FOur prior experience with the use of MCMC methods in nonlinear dynamic models highlights the risk that model mis-specifications may lead to confidence intervals that are too tight. We therefore downscale the likelihood function to expand the confidence intervals (details in the online appendix) to err on the side of caution in assessing uncertainties and structural nuances not explicitly modeled.

## 3. Results

We first provide results from an out of sample prediction exercise intended to build additional confidence in our method, then discuss the detailed results based on full sample.

### 3.1. Out of sample prediction test

Our data are limited (three data series for 30 days plus a handful of data points for other variables) and thus proper out of sample prediction test is limited in its scope. Nevertheless, we calibrate the model with the first 15 days of data using the methods discussed above, and test the model’s ability in replicating the rest of the data points. Figure 2 shows the results, comparing simulation outcomes (median and 90% confidence interval) with data.

**Figure 2.**
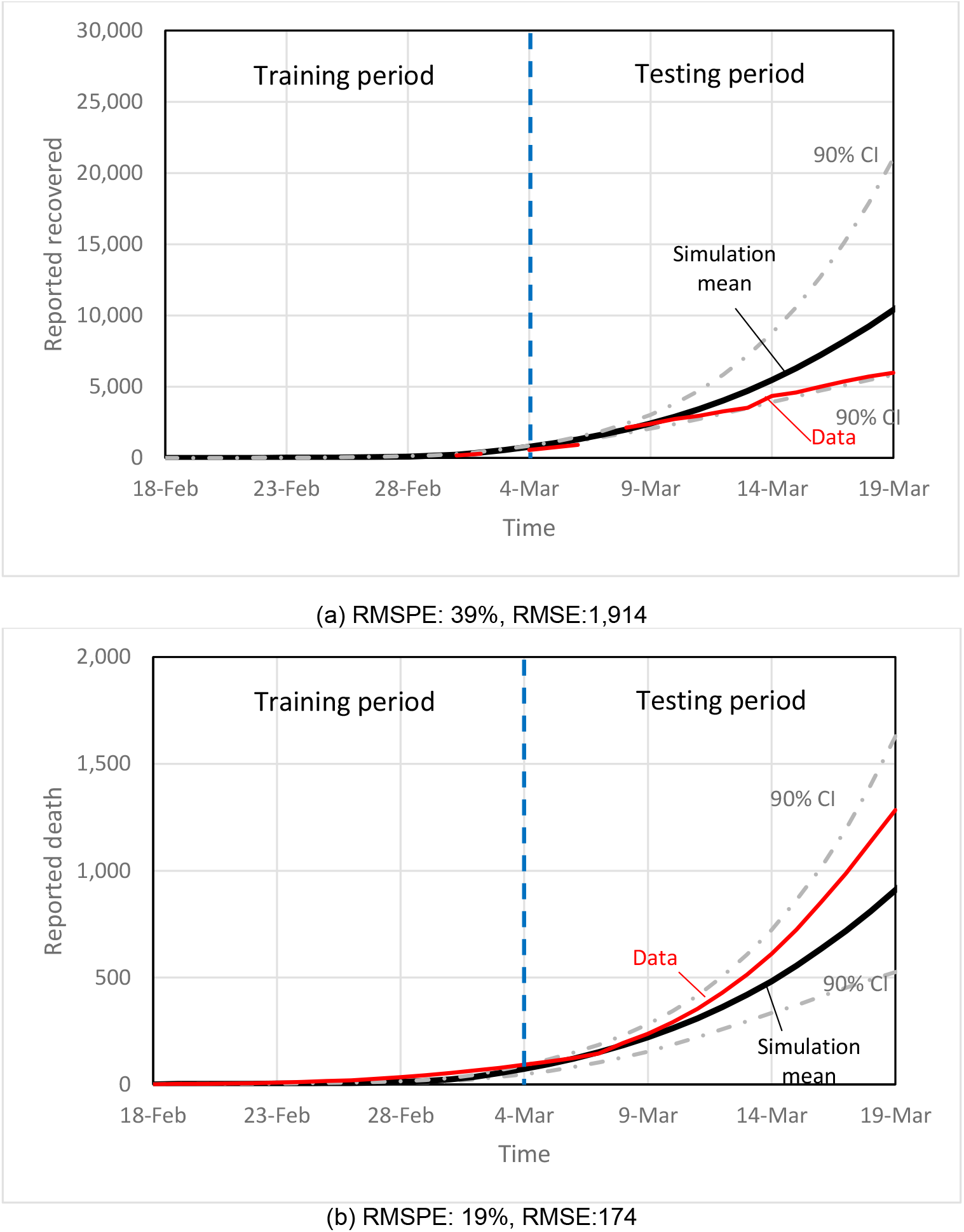
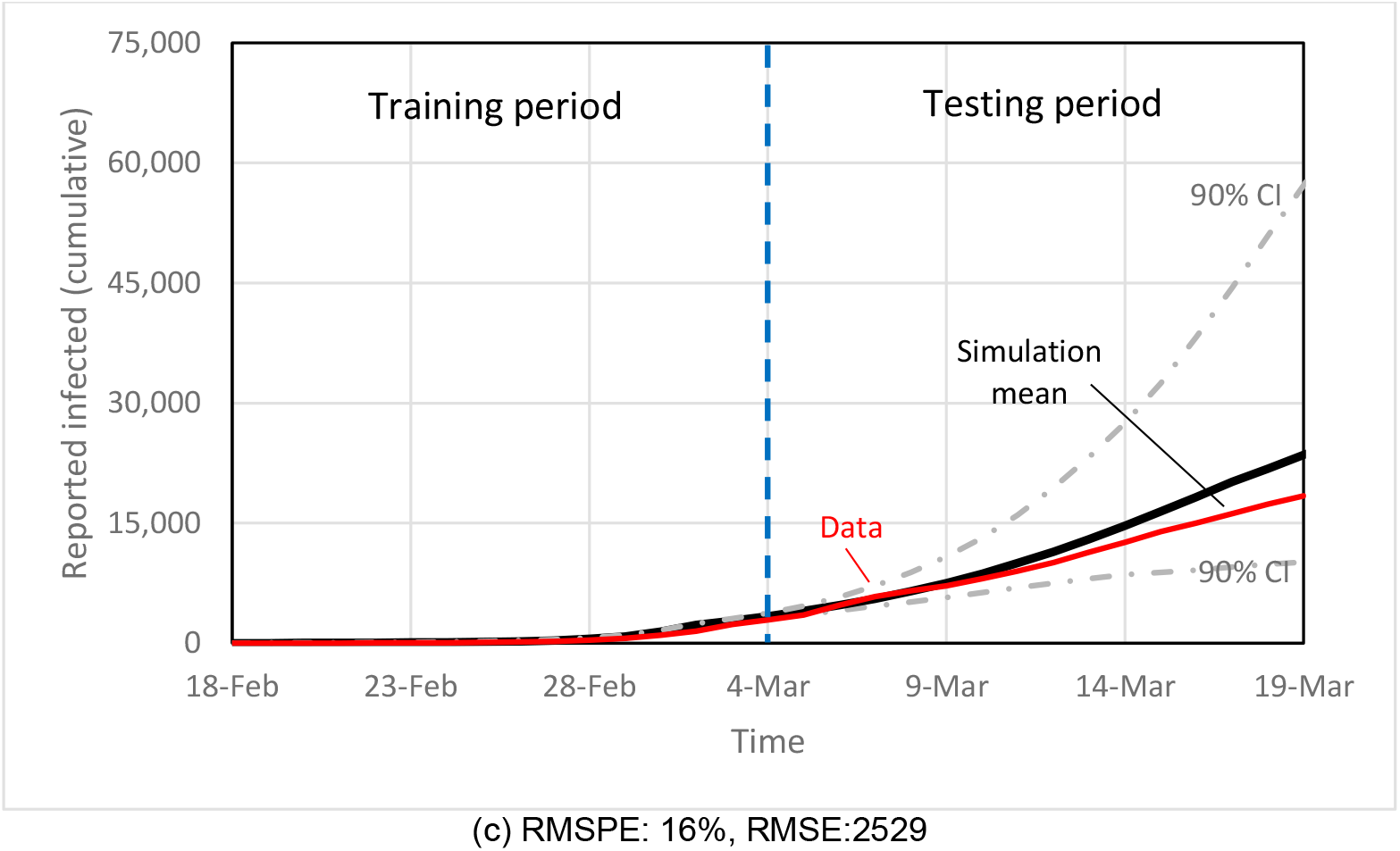
Comparison of the simulation predications of reports (mean and 90% confidence interval (CI)) with data. The model is calibrated for the first 15 days of data and testing against the last 15 days of data. Note: RMSPE: Root mean square percentage error, RMSE: Root mean square error.

The model performs better in replicating cumulative cases of infection and death than recovered. While any model that may create exponential growth trend might perform reasonably under this tests, our model is constrained by many structural features of contagion and behavioral response dynamics, and the match is despite those constraints. In the next section, we recalibrate the model with the entire data of 30 days.

### 3.2. Parameter estimation

We follow the procedure described in the modeling section and calibrate our model using multiple data sources. The results from MCMC is a joint distribution of parameter values for 11 estimated parameters through calibration. After about 2 million simulation runs, we received about 1.6 million acceptable points from posterior joint distribution of parameters, and used top 90% of the distribution of payoff values. We randomly selected 3% of those samples and used those to simulate the trajectories for various model variables. Figure 3-a compares simulation results for official confirmed cases of death, recovered, and cumulative cases of infection compared with the data. RMSPE values of simulation for the last 10 days of reported recovered, death, and cumulative infected are 16%, 26%, and 5%, and RMSE’s are 877, 116, and 794. Simulation runs almost perfectly correlate with data. Table A1 in the Appendix reports estimated parameters along with those adopted from the literature. Figure 3-b shows how simulation runs replicate unofficial statistics including the number of infected travelers diagnosed before air travel patterns changed substantially, and medical community’s counts of cumulative death reported by media sources.

**Figure 3.**
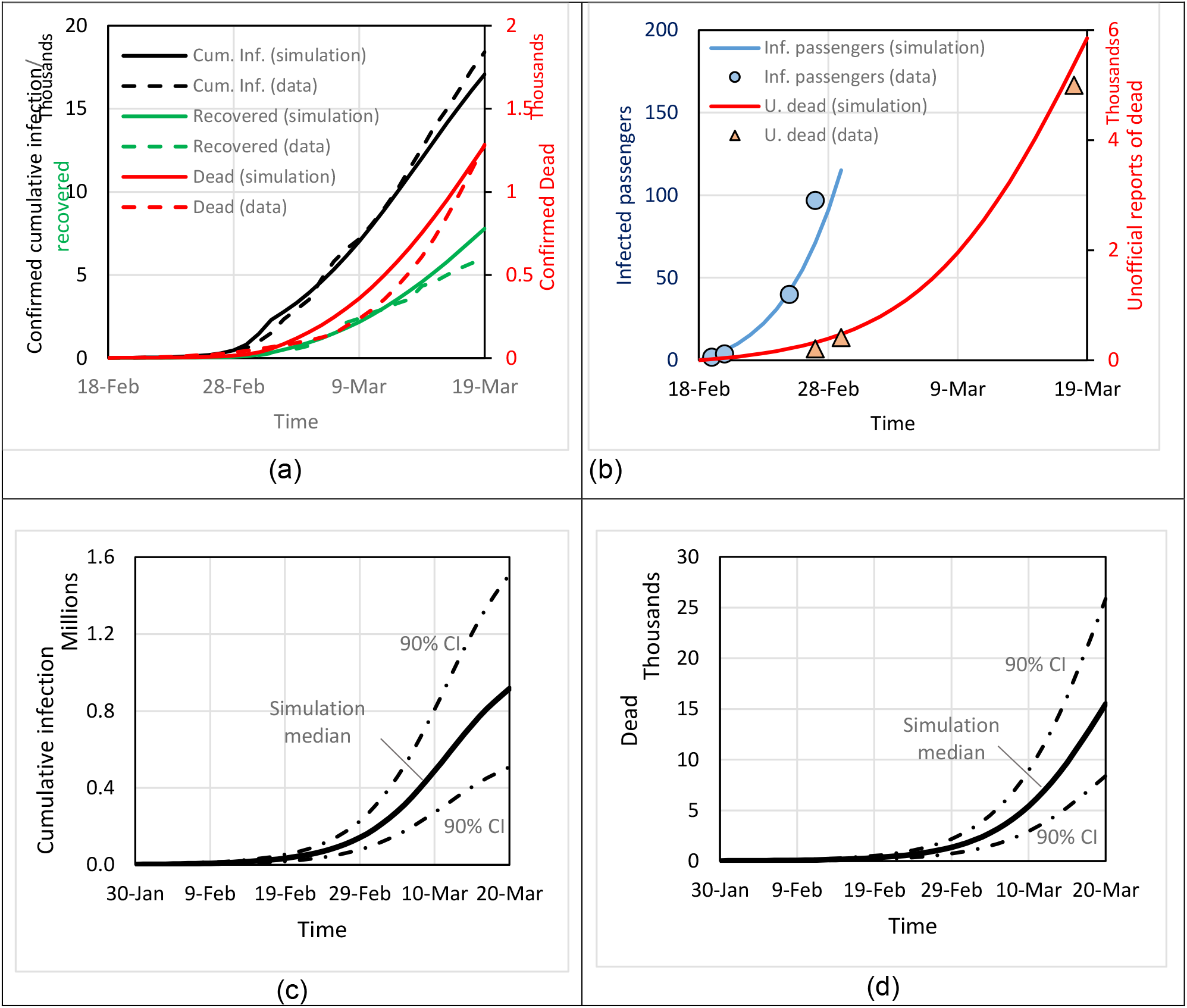
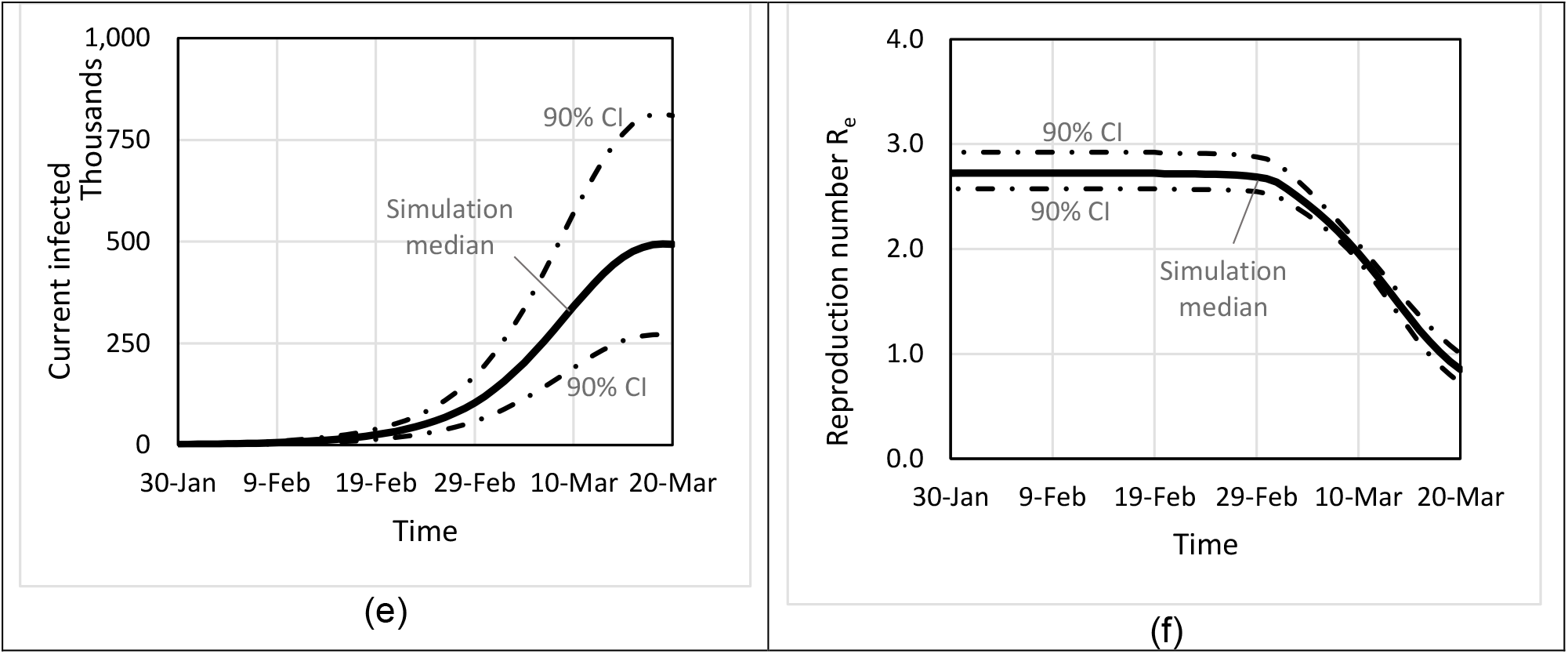
Base run simulations: Replication of data for confirmed cases (a), unofficial reports and infected passengers (b); simulation-based estimation of cumulative infected (c), death (d), current infected population (e), and reproduction number. Panels c-e include median and 90% confidence intervals. Note: Cum Inf.: official reports for cumulative cases of infected;, Inf. Passengers: Infected passengers diagnosed at international airports; U. dead: Unofficial reports about cases on death; CI: confidence interval.

### 3.3. Status of COVID-19 in Iran as of March 20 2020

The simulation results reported in Figure 2 include simulation-based estimation of cumulative number of infected (panel c), cumulative number of death (panel d), and current number of infected (panel e) until March 20, 2020, starting from December 31^st^, 2019 (day 0 in simulation). As discussed in detail in the modeling assumptions (above) and limitations (below), there are several structural uncertainties that can affect our estimations. Some of those uncertainties are quantified further in section 3.4. With these sources of uncertainty in mind, we find total infected cases at the time of this writing may be closer to 916,000, than the reported 19,000. Cumulative deaths by March 20th, over 15000, may also be an order of magnitude higher than official statistics and might have almost tripled in the last 10 days of the analysis. There is also much uncertainty in these numbers: our baseline estimate for cumulative infected to date may be as low as 508,000 and as high as 1.5 million. Similarly cumulative deaths is between 8400 and 25,800.

The wide confidence intervals are partly due to the nature of the data: not having found any data on the rate of testing in Iran it is very hard to fully know the underlying diffusion patterns based only on the formal statistics. The travel and informal estimates of death partially address these limitations, but much uncertainty remains. Our conservative assumption on downscaling the likelihood function may also have contributed to this wide interval. Nevertheless, even our lower bound are 26 times the official statistics for total infections and 6 times larger than the death statistics.

There are some glimmers of hope in our estimates as well. The drop in contact rate in response to perceived risk, which we have captured in our model, might have reduced R_e_ significantly, to just below 1 as of late March (panel f). That reduction, if confirmed by data in the coming days, would help bring down the number of new cases. Thus, Figure 3-e shows the number of currently infected cases may well be reaching its peak at the end of the simulation. Note that this number is smaller than cumulative cases to date (panel c).

The epidemic seems to have raged with a reproduction number close to 2.72 (90% CI: 2.57-2.92 CI) for more than six weeks before the behavioral and policy interventions have slowed down the diffusion rate starting in late February. Some of this drop may also be attributed to weather, but current research is mixed on the impact of weather (Luo, Majumder et al. 2020, Wang, Jiang et al. 2020) so we did not include it in our estimation (but would explore it in forward projection in 3.5). Those factors, we estimate, have already brought down the aggregate contact rate to 26% (90% CI: 22%-31%) of pre-epidemic levels. This number may just succeed in bringing down the reproduction number below 1, which is the necessary condition for containment of the epidemic.

Whereas the case fatality rate in Iran, based on official statistics until March 20^th^, is 18.4%, our estimated case fatality rate for the Iranian population is 3.7 (0.4)% (standard errors for estimates are in parentheses). This estimate is consistent with, and slightly higher than, the 2.3% reported by Wu and McGoogan (2020) based on tracing confirmed cases in China. If correct, the magnitude of epidemic in Iran would be much larger than that in China so far, and limits to healthcare services may explain the increased death rate. This finding is reinforced by the limited coverage of testing in Iran that is indicated in our estimation. Many cases go unconfirmed, and at its maximum, formal testing is covering only 2.5 (0.9)% of infections and 26 (17)% of deaths. These estimates are consistent with qualitative media reports from Iran on very limited availability of testing, the doctors’ need for getting authorization before conducting tests, and the multi-day delays in receiving test results that render them ineffective in the clinical decision making process. We estimate that hospital sources of news organizations who have offered alternative versions of actual death statistics in Iran have had a much wider coverage of true death statistics (42 (15)% of true values). Their count offers a lower bound for true deaths, is also close to our lower confidence bound, and suggests that the true number of cumulative cases, even with a case fatality rate as high as 3.7%, would not be lower than 380,000.

Nevertheless, we estimate that even those medical community reports offer coverage significantly below 100%, consistent with the less-than-perfect coverage of hospital reports by media sources, and anecdotal evidence that hospital system has been overwhelmed in most hot spots of the disease and many patients have died at home and been buried with no testing or proper association of cause of death to COVID-19.

### 3.4 Sensitivity to disease parameters

The model used prior estimates from the literature for three important parameters: the total duration of illness (d=14 days), the asymptomatic period (τ=4 days), and the fractional infectivity of the asymptomatic period (θ=0.25). We assess our projections’ sensitivity to these structural assumptions by re-estimating the model for different values of each parameter. In this analysis we find a new set of best-fitting parameters given each different assumption on disease parameters. In Table 1 results are summarized as percentage changes in key projections (cumulative infections and cumulative death until March 20^th^, 2020) compared to baseline best-fitting parameters.

**Table 1.** Impact of alternative disease parameters.

| | $\theta = 0.4$ | $\theta = 0.15$ | $\tau = 5$ | $\tau = 3$ | $d = 16$ | $d = 12$ |
| --- | --- | --- | --- | --- | --- | --- |
| Cumulative Infection | -22.0% | 44.0% | 18.2% | -4.6% | 1.5% | 21.3% |
| Cumulative Death | -21.2% | 44.0% | 18.5% | -4.9% | -7.3% | 34.7% |

Note: Each row reports percentage change in cumulative infection and death compared to baseline given alternative assumptions on fractional infectivity of asymptomatic period ( $\theta$ ), asymptomatic period ( $\tau$ ), and duration of illness ( $d$ ).

Results suggest these structural parameters are important in identifying the true magnitude of the epidemic. Specifically, a smaller fractional infectivity during asymptomatic period (τ), a longer asymptomatic period (τ), or a shorter duration of illness all significantly increase the projections for total cases and deaths. Reductions in the estimates are less significant when parameters move in the opposite direction, though a very high level of θ would bring down the estimated cumulative cases notably. Current evidence for transmission during asymptomatic period is rather limited, and thus our baseline projections may be somewhat conservative. Nevertheless, given our conservative treatment of the confidence intervals all these baseline projections with different disease parameters remain within the 90% bounds of our baseline projections.

### 3.5 Future trajectory of the critical measures

We run the model until July 1st of 2020 under six scenarios (three alternative assumptions about the impact of seasonality times two policy measures). Specifically, our scenarios about seasonality include no effect (status quo), moderate effect (infectivity of the virus decreases linearly from April 1^st^ and halves by June 1^st^, then stays the same for the rest of the simulation), and very strong mitigating effect (infectivity of the virus decreases from April 1^st^ to a quarter of its base value by June 1^st^, then stays the same for the rest of the simulation). For policy, our focus is on contact rate, and we include two conditions of status quo and aggressive efforts to decrease contact rate by half what it would be otherwise. These six conditions provide intuitions for a potentially wide range of cases. Both intervention and weather impacts are assumed to include rather strong options, which is to offer a feel for the range of possibilities, but not necessarily be representative of most likely scenarios. Moreover, we also note that the wide cone of uncertainty in the baseline simulations will continue and expand in future projections.

Keeping the uncertainty considerations in mind (but not graphed due to clutter), Figure 4 shows results based on best-fit parameters. If our best case scenario on the reduction of contacts and benefits of seasonality for containment hold, the number of infected cases will peak soon at approximately 494,000 (90% CI: 274K-813K), and will go down later on. This optimistic scenario will still lead to over a million infections and some 58,000 deaths (90% CI: 32K-97K) by the end of June. The less aggressive scenarios point to continued spread of the epidemic. A reduction in contagion is realized only when reproduction number remains below one as a result of natural contact reduction in response to recent deaths, government interventions that reduce contacts, and seasonality. Among these three, the first (behavioral response) is endogenous to recent death rate. When death rate comes under control (due to the combination of all those factors), our model assumes the contract rate rebounds, weakening the first channel. Economic pressures, normalization of death, and behavioral modeling after the more risk seeking (or those already recovered and thus presumably immune) could all weaken the behavioral response when the perceived risks have faded. The increased contact would then, with a delay, bring up the death rate close to levels sustaining a reproduction number around or slightly above one. This creates a strong attractor in the dynamics where in steady state contact rate is high enough to sustain the contagion but not so high to lead to the rapid infection of all the population. Therefore it is only with strong weather and/or government interventions that reduced reproduction number brings the epidemic under control. This dynamic offers a cautionary tale against declaring victory early in the fight against the epidemic. However the actual magnitude of such rebound effect is not known in this setting and our data offers no guidance on the relevant parameters. Therefore these results are only qualitatively suggestive but not quantitatively reliable.

**Figure 4.**
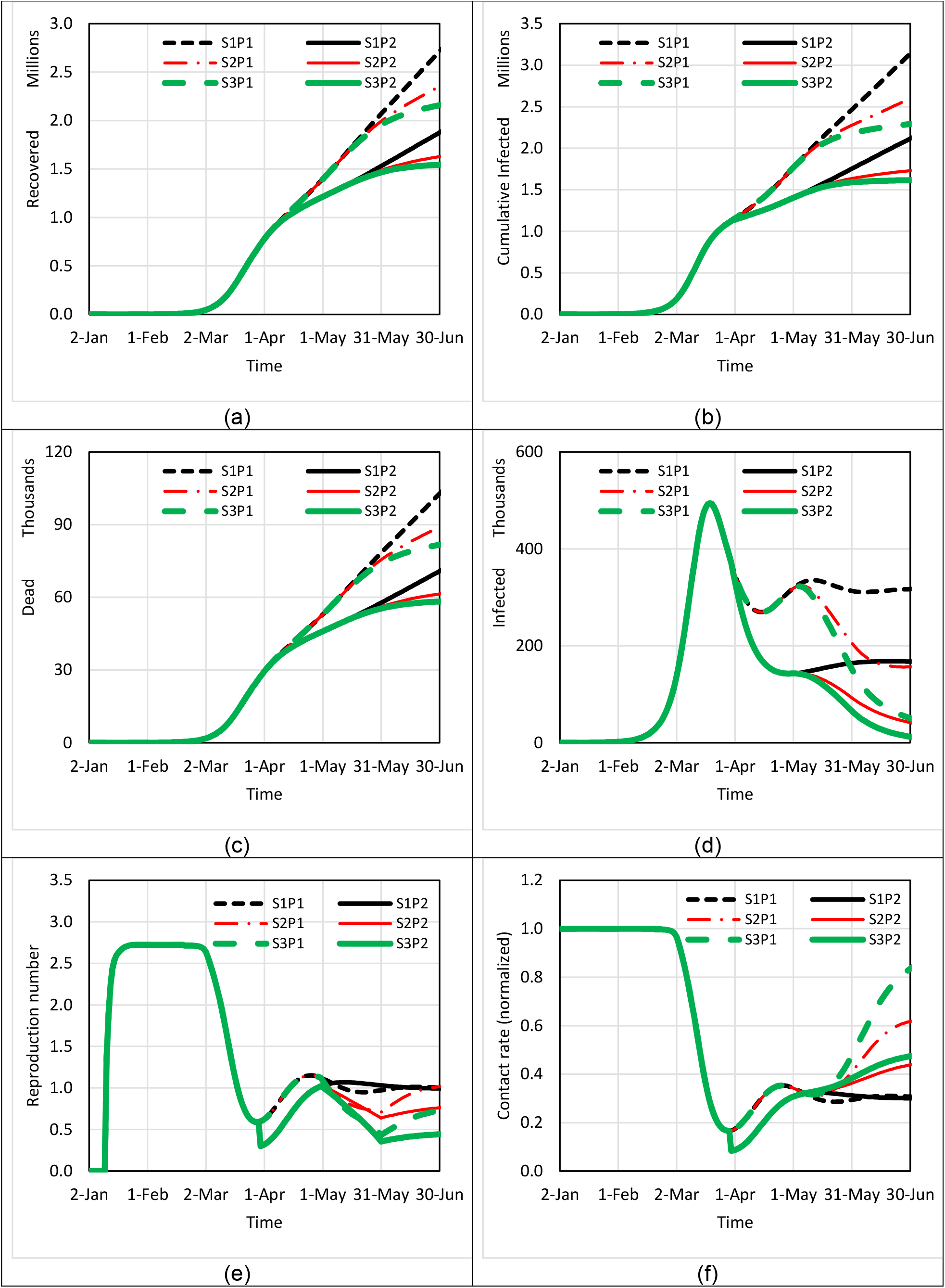
Simulation-based estimation of the trajectory of cumulative recovered (a), death (b), cumulative infected (c), current infected (d), Reproduction number (e) and Contact rate (f), until the end of June 2020, under 6 conditions: (Three seasonality effect conditions (S1-S3) X two policy effects (P1 and P2)). Note: Each graph shows the median value of than 3000 simulation runs. 90% CI would give a wider range of results. The reproduction number starts from zero given that the disease have slightly different starting days at different simulations, and then reaches its initial steady state value.

## 4. Discussion

In this paper we provided a more sobering picture of the COVID-19 outbreak in Iran using a dynamic model that goes beyond official statistics. Integrating data from various sources we suspect the current official statistics are at least an order of magnitude below actual spread of the epidemic, and even in optimistic scenarios the burden of disease will be large and lasting for many months. Implementing, and sustaining, strong policies that target social distancing offers the main hope for containing the epidemic.

The gap between official statistics and our estimates may be due to various complications in measurement as well as policy choices. The availability of testing infrastructure has been a major bottleneck for detecting cases in Iran. Citizens with suspicious symptoms have had no easy way for getting tested; most tests are conducted on hospitalized patients, require specific authorization, and are processed in a few centralized labs adding significant delays to the process. Not only mild cases are missed, but also many critical patients have been unable to access the care they need due to hospital congestion. The fact that majority of cases have mild symptoms similar to flu (Wu and McGoogan 2020) adds to the risk of under-counting even if the testing capacity was ample. In fact many mild or moderately sick patients may have preferred to stay home than risk being infected upon visiting a congested hospital, further reducing demand for testing.

Many of the insights form this analysis are applicable to the spread of the disease in other regions. Specifically, many countries have low test rates which may be hiding the magnitude of the epidemic and increasing the risk of unpleasant surprises for policy makers and the public.

This paper is based on modeling-in-real-time of an emerging outbreak. The problem is rapidly changing, and timely estimation is critical for the purpose of our analysis. Therefore we have made several simplifying assumptions that readers should be aware of in interpreting our results. First, consistent with prior findings (Rahmandad and Sterman 2008) we have focused on capturing behavioral feedbacks in contacts and testing and have adopted a formal estimation process, but have done so at the cost of abstracting away from much detail complexity and heterogeneity in populations and risks. Second, we have left out explicit treatment of healthcare resources and their impact on the burden of disease. Third the model is scoped only around Iran -- we ignore potential effects of global spread of the disease on Iran, including the risk of reintroduction of cases in future. Forth, we assumed that the recovered population are immune during our simulation time period of six months. Fifth, we ignore mutations in the virus which may change its contagiousness and case fatality rate. Despite these limits we hope the paper offers a more accurate assessment of the risks and scope of the epidemic for Iran and beyond. We also hope other researchers build on this work using the publicly available data, models, and replication instructions provided in the online appendix.

## Data Availability

Publicly available data are used and all are reported in the paper or supplementary material.

https://osf.io/v2d7q/?view_only=1a92d113520243b6985614a1ec17315c

## Acknowledgments

We are thankful to Mohammad Akbarpour, Narges Doratoltaj, Babak Heydari, Hamed Ghoddusi, and Tse Yang Lim for their thoughtful feedback on various drafts of this paper.

## Appendix A summary of the article in Persian

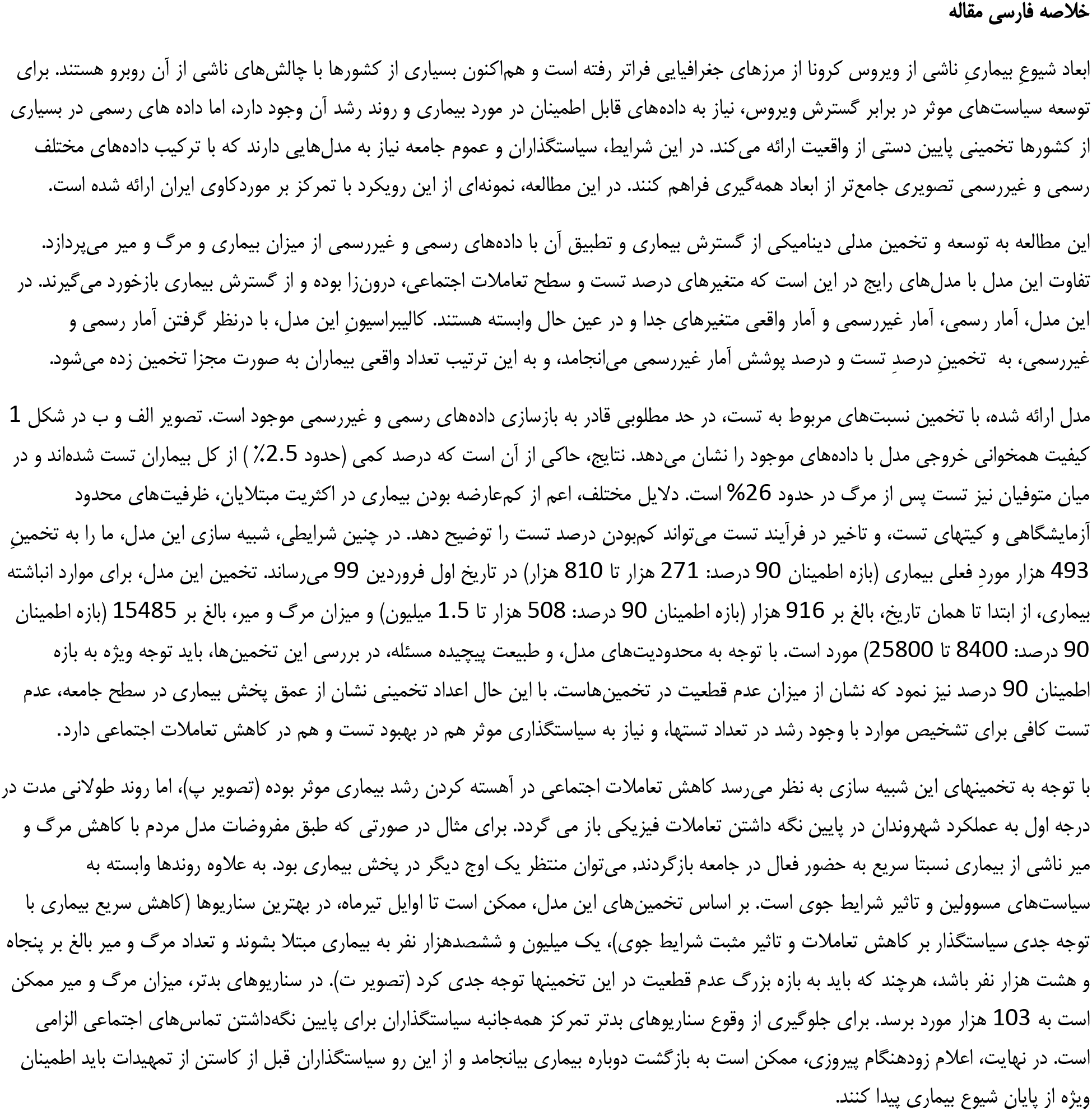
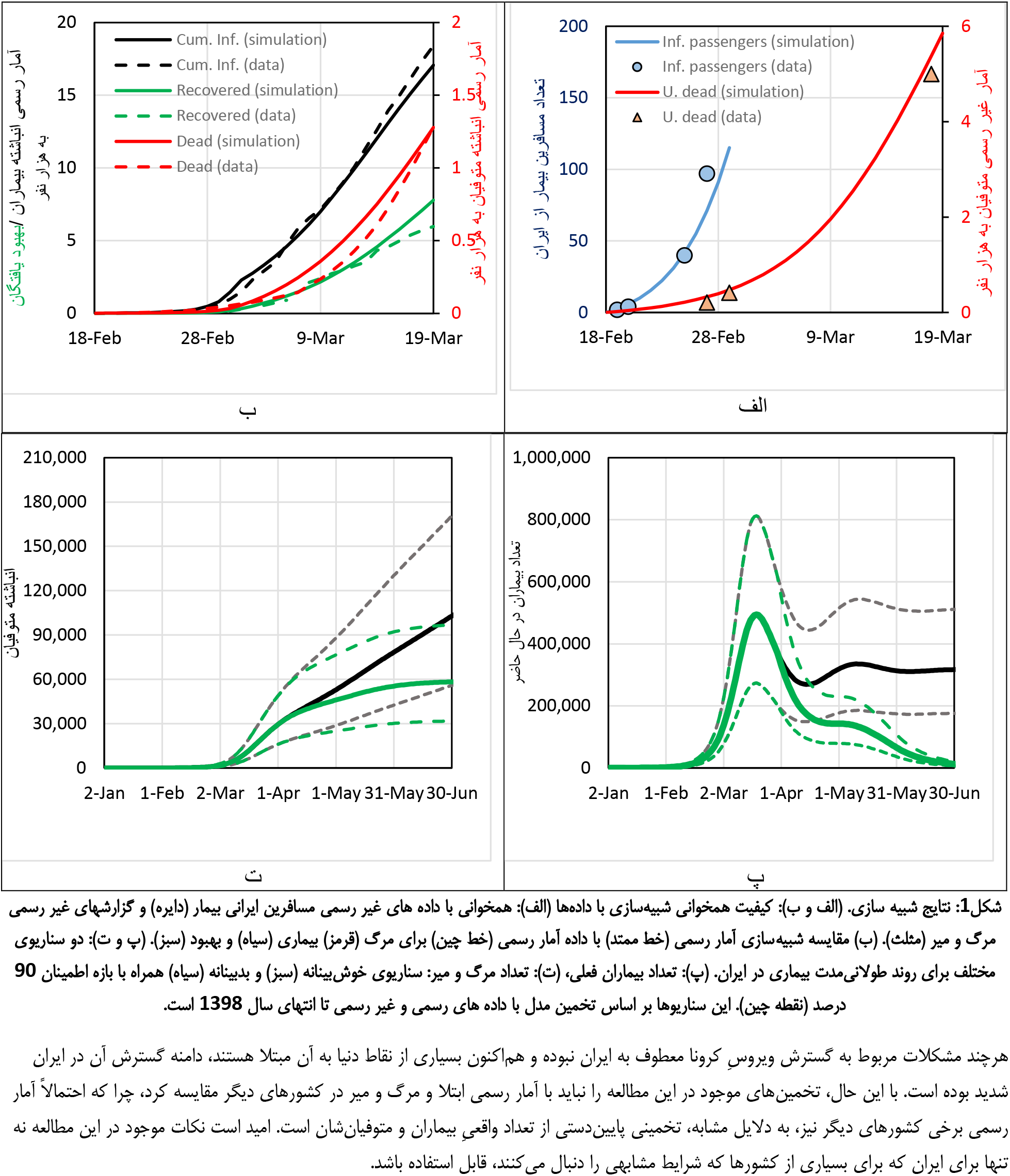

